# Prospective Multicenter External Validation of the BIDIAP Index for the Diagnosis of Pediatric Acute Appendicitis

**DOI:** 10.64898/2025.12.31.25343255

**Authors:** Javier Arredondo Montero, Andrea Herreras Martínez, Luis Rello Varas, Alicia Escudero Villafañe, Marina Iglesias Oricheta, Maria del Mar Larrea Ortiz-Quintana, Lucía Fernández Rodríguez, Pablo Aguado Roncero, María Carmen Campos Calleja, Ricardo Díez, Samuel Sáez Álvarez, Carmen Ruiz de la Cuesta Martín, Carlos Delgado Miguel, Rafael Fernández Atuan

## Abstract

**Introduction:** Pediatric acute appendicitis (PAA) remains challenging to diagnose despite existing diagnostic scores. The BIDIAP index is a three-item diagnostic tool with very high discriminative performance in a derivation cohort. This study aimed to prospectively and externally validate the BIDIAP index in a multicenter pediatric population.

**Material and Methods:** We conducted a prospective, multicenter observational study across four tertiary pediatric centers, enrolling children presenting with suspected PAA. Two groups were analyzed: patients with histopathologically confirmed PAA and patients in whom appendicitis was confidently excluded after diagnostic work-up, classified as non-surgical abdominal pain (NSAP). The BIDIAP index was applied using a predefined cutoff (≥ 4 points), and diagnostic performance was assessed using ROC analysis, calibration metrics, and decision curve analysis (DCA).

**Results:** A total of 644 patients meeting the prespecified analytical criteria were included in the primary analysis. The BIDIAP index demonstrated excellent diagnostic performance, with an area under the ROC curve of 0.93 (95% CI, 0.92–0.95). The calibration slope was 1.00, and the intercept was close to zero, indicating close agreement between predicted and observed risks. At the prespecified cutoff value of ≥ 4 points, the BIDIAP index achieved a sensitivity of 90.5% and a specificity of 81.6%. DCA showed a positive net clinical benefit of the BIDIAP index over treat-all and treat-none strategies across the full range of clinically relevant threshold probabilities.

**Conclusions:** The BIDIAP index demonstrated excellent diagnostic performance for PAA. Its simplicity, based on only three items, and its potential applicability even when the appendix is not visualized on ultrasonography make the BIDIAP index a promising tool for supporting clinical decision-making in routine pediatric emergency practice.

## Introduction

Pediatric acute appendicitis (PAA) is the most common surgical emergency in childhood worldwide [1]. Its diagnosis is based on integrating clinical assessment, peripheral blood analysis, and imaging studies. Despite this multimodal diagnostic strategy, clinically relevant diagnostic errors remain frequent, contributing to preventable morbidity and worse outcomes [2].

Multivariable diagnostic scores represent a helpful adjunct in the evaluation of suspected PAA, aiming to standardize clinical assessment and optimize diagnostic performance. Among these, the Alvarado score was one of the earliest and most widely adopted tools, originally developed and validated primarily in adult populations [3]. However, its performance in children has been inconsistent, prompting concerns regarding limited discriminative ability and reduced applicability in pediatric settings [4,5]. Consequently, several pediatric-specific risk stratification tools have been developed, including the Pediatric Appendicitis Score (PAS) [4,6,7] and the Pediatric Appendicitis Risk Calculator (pARC) [8,9], which incorporate population-appropriate clinical and laboratory variables to better reflect the presentation of acute appendicitis in children and to improve diagnostic accuracy. Nevertheless, both PAS and pARC are multi-item instruments that require integrating several clinical and laboratory variables, which may limit their practical applicability in time-pressured settings. In particular, the PAS comprises eight components. In contrast, the pARC incorporates nine variables, including modifiers such as care setting, potentially limiting rapid bedside calculation in time-pressured Emergency Department environments. Consequently, despite generally good diagnostic performance on external validation [4,6–9], their use may be time-consuming in busy emergency department environments.

Previous studies have reported sociodemographic disparities in the diagnosis and outcomes of PAA [10], including differences related to age, sex, and socioeconomic context, which may affect time to diagnosis and complication rates. Such inequalities highlight the importance of diagnostic tools that are simple, standardized, and applicable across diverse clinical settings.

In 2023, our research group developed a novel diagnostic index, termed the BIDIAP index, composed of three readily available items. The index was derived from a prospective single-center cohort and identified ultrasonographic appendiceal caliber, systemic immune-inflammation index (SII), and peritoneal irritation as key predictors of PAA [11]. In the derivation cohort, the BIDIAP index demonstrated very high discriminative performance, with an area under the receiver operating characteristic curve of 0.97. However, as this performance was obtained on a derivation dataset, the possibility of overfitting cannot be ruled out. Consequently, the primary objective of the present study was to perform a prospective multicenter external validation of the BIDIAP index to assess its diagnostic performance and generalizability in routine clinical practice.

## Material and methods

### Study Design

Building upon the development of the BIDIAP index, BIDIAP-2 (Biomarkers for the DIagnosis of Appendicitis in Pediatrics 2) was a prospective, observational, multicenter study conducted across four Spanish tertiary referral centers between March 2024 and April 2025. Its primary objective was to validate the BIDIAP index for diagnosing pediatric acute appendicitis (PAA). The study consecutively enrolled pediatric patients (0–18 years) presenting with a formal clinical suspicion of PAA. Participants were subsequently stratified into two groups: (1) those with histopathologically confirmed PAA (PAA group) and (2) those in whom appendicitis was reliably excluded (non-surgical abdominal pain, NSAP group). Histopathological examination of the surgical specimen served as the reference standard for diagnosing PAA. In the NSAP group, PAA was excluded based on clinical assessment, identification of an alternative diagnosis when applicable, and structured follow-up confirming the absence of subsequent appendicitis. Losses to follow-up were recorded. Patients who underwent appendectomy but showed no histopathological evidence of acute appendicitis (negative appendectomy) were classified within the NSAP group.

Approval from the institutional review board was obtained before study initiation. Before enrollment, written informed consent was obtained from the parents or legal guardians of all patients. All clinical and sociodemographic data were anonymized in accordance with applicable data protection regulations. The study was conducted in accordance with the principles of the Declaration of Helsinki (2013 revision) and reported according to the STARD 2015 (Standards for Reporting Diagnostic Accuracy Studies) guidelines [12] and the TRIPOD+AI (Transparent Reporting of a multivariable prediction model for Individual Prognosis Or Diagnosis) statement for the external validation of prediction models [13] (Supplementary Files 1 and 2). Supplementary File 3 details inclusion and exclusion criteria.

### Sample Collection

In all patients, a peripheral venous blood sample was obtained upon arrival at the Pediatric Emergency Department as part of the initial diagnostic workup. Specifically, blood was drawn into three separate vacuum tubes: a K2-EDTA tube for the complete blood count (CBC) and calculation of the SII, a sodium citrate tube for coagulation studies, and a serum separator tube (SST) with clot activator for biochemical analysis. For each tube, 2.5-4 mL of blood was collected. Samples were processed following the specific protocols of each Clinical Analysis Department and in strict accordance with the manufacturers’ operating instructions for the automated analyzers.

### Data Collection, Storage, And Processing

All study data were collected, stored, and processed securely using encrypted systems with restricted access. Access to the data was limited to the study investigators, and the data were kept anonymized at all times, in compliance with applicable data protection regulations.

### Systemic-Immune Index Calculation

For the calculation of the SII, the absolute neutrophil count (ANC), absolute platelet count (APC), and absolute lymphocyte count (ALC) were extracted from the blood count. The SII was calculated using the formula: SII = (ANC × APC) / ALC, as previously described in the literature. All hematological values (ANC, APC, ALC) were expressed in International System units (×10 /L) while the SII is an index and therefore is dimensionless.

### Ultrasonographic Measurements and Data Collection

The attending radiologist on duty performed a diagnostic ultrasound examination on all patients at the time of emergency evaluation. Data collection was conducted using a standardized template developed prior to study initiation.

### Calculation of the BIDIAP Index

The BIDIAP index was calculated based on the three independent predictors identified in the original derivation cohort. These variables were dichotomized using optimal cut-off points. Specifically, the index assigns 4 points for an appendiceal caliber ≥ 6.9 mm, 3 points for an SII ≥ 890, and 2 points for peritoneal irritation (PI).

The total score is the sum of these components, ranging from 0 to 9 points. In the original study, a cut-off value of 4 points was established as the optimal threshold for diagnosing pediatric acute appendicitis (PAA). Based on this threshold, a diagnosis of PAA is confirmed if the appendiceal caliber is ≥ 6.9 mm (scoring 4 points) or if the patient presents with both an SII ≥ 890 and peritoneal irritation (scoring 3 + 2 = 5 points).

For the present study, the BIDIAP index was calculated for all included patients, maintaining the predefined cut-off of ≥ 4 points as the primary validation threshold. Additionally, alternative cut-off points were explored to assess their diagnostic performance and clinical utility in this multicenter cohort.

Although PI was analyzed descriptively as a three-level variable, cases classified as unclear were coded as negative (0 points) for inclusion in the primary analysis. Likewise, when ultrasonography was performed, but the appendiceal caliber was not reported, the corresponding index component was assigned 0 points.

### Statistical Analysis

#### Sample Size

Before study initiation, the sample size was calculated to ensure adequate power to externally validate the discriminative performance of the BIDIAP Index against a conservative null hypothesis reflecting commonly used pediatric appendicitis scores. The null hypothesis was set at an AUC = 0.85 (representative of widely used pediatric appendicitis scores), versus an alternative hypothesis of AUC = 0.90 for BIDIAP, assuming a more conservative performance than that reported in the derivation phase (AUC = 0.97) to reflect an external validation scenario. Using the Hanley–McNeil methodology for ROC analysis [14,15], with a two-sided significance level of 0.05 and 80% statistical power, and assuming an estimated prevalence of 50% of pediatric acute appendicitis in the study population (patients with formal clinical suspicion), the required sample size was 322 patients (161 per group). Based on missing data observed in the original derivation study, where incomplete appendiceal caliber measurements led to patient losses, an additional safety margin was applied to the estimated sample size. Accordingly, the target recruitment was increased by approximately 10–15% to account for potential missing data, resulting in a final planned sample size of approximately 360–370 patients.

#### Analytical Sample Definition and Missing Data Handling

The primary analysis included all patients with available data for all components required to compute the BIDIAP score, defined *a priori* as: (1) complete blood count enabling calculation of the SII, (2) a documented assessment of PI, and (3) a performed ultrasonographic examination. When ultrasonography was performed but the appendiceal caliber was not reported, this component was coded as 0 points per a prespecified operational rule. No statistical imputation of missing data was performed. Patients lacking any of the required components for score calculation were excluded from the primary analysis.

#### Statistical Analysis for Group Comparisons

Statistical assumptions were verified before test selection. The normality of continuous variables was assessed using the Shapiro–Wilk test and visual inspection of histograms. Homogeneity of variances between groups was evaluated using the Brown–Forsythe test (median-centered). Depending on distributional characteristics, either parametric tests (Student’s t-test) or non-parametric alternatives (Mann–Whitney U test) were applied. Categorical variables were compared using the Chi-square test or Fisher’s exact test when expected cell counts were below 5. Given the descriptive nature of baseline comparisons, p-values were not adjusted for multiple testing.

#### Data Visualization and Graphical Analysis

Raincloud plots were used to visualize the distribution of selected continuous variables across groups. Medians and interquartile ranges were reported to summarize central tendency and dispersion.

#### Receiver Operating Characteristic Curve Analysis

Receiver operating characteristic (ROC) curve analysis was performed to evaluate the discriminative performance of continuous variables. The area under the ROC curve (AUC) was used as a global measure of diagnostic accuracy. As a secondary robustness check, 10-fold stratified cross-validation was performed to assess the stability of discrimination across subsamples of the validation cohort. Cross-validation was not used for model fitting or optimization.

Additionally, non-parametric bootstrap resampling (1,000 iterations, percentile method) was used to estimate the 95% confidence intervals for the AUC. Cross-validation and bootstrap were used in a complementary manner to assess the stability of discrimination estimates and population-level variability. AUCs were compared using the DeLong test for correlated ROC curves.

#### Calibration

Calibration of the BIDIAP index was evaluated by comparing predicted probabilities with observed outcome frequencies across deciles of risk using calibration plots. To obtain predicted risks for calibration, a logistic regression model was fitted using the BIDIAP index as the sole predictor, and the resulting predicted probabilities were used for subsequent analyses. Overall model performance was further assessed using the Brier score as a measure of prediction accuracy, with lower values indicating better performance. In addition, calibration slope and calibration intercept were estimated by regressing the observed outcome on the logit of the predicted risk; a slope close to 1 and an intercept close to 0 were considered indicative of good calibration across the multicenter cohort.

#### Decision Curve Analysis

The clinical utility of the BIDIAP index was explored through decision curve analysis (DCA) [16], which quantified the net benefit of each strategy across a range of threshold probabilities, compared to default strategies of treating all or treating none.

#### Sensitivity Analyses

Two prespecified sensitivity analyses were performed to assess the robustness of the BIDIAP index under alternative, clinically plausible assumptions.

First, because the interpretation of peritoneal irritation (PI) may be uncertain in a subset of patients, a sensitivity analysis was conducted in which cases classified as unclear for PI were considered positive. In contrast, in the primary analysis, they were coded as negative.

Second, to specifically evaluate the impact of ultrasonographic availability of the highest-weighted score component, a sensitivity analysis was performed, restricting the analysis to patients in whom the appendiceal diameter was directly measured on ultrasonography. In both analyses, the original score structure, variable definitions, and prespecified cutoff values were maintained, and diagnostic performance, calibration, and clinical utility were reassessed using the same statistical framework as in the primary analysis.

#### Statistical Significance Level and Software

All tests were two-tailed, and p-values < 0.05 were considered statistically significant. Statistical analyses were performed using Stata version 19.0 (StataCorp LLC, College Station, TX, USA) and R version 4.5.1 (R Foundation for Statistical Computing, Vienna, Austria).

## Results

### Study Population

A total of 725 children evaluated for suspected acute appendicitis were prospectively enrolled during the study period. After excluding patients with missing key diagnostic data, lack of informed consent, incorrect inclusion criteria, or inadequate reference standard (n = 6), 719 patients were retained in the final cohort. This cohort comprised 305 patients in whom PAA was confidently excluded (classified as NSAP) and 414 patients with histopathologically confirmed acute appendicitis (PAA). Of the patients included in the NSAP group, 23 corresponded to cases of negative appendectomy, in which surgical exploration was performed but histopathological examination excluded PAA. Data required to compute the BIDIAP score according to the prespecified operational criteria were available for 644 patients. For this definition, data completeness required that an ultrasonographic examination had been performed, regardless of whether the appendix was directly visualized, in keeping with the predefined operational rules of the BIDIAP index. Patient flow, exclusions, and reference standard assignment are detailed in Supplementary File 4.

### Baseline Sociodemographic and Clinical Characteristics

At the sociodemographic level, a statistically significant difference in sex distribution was observed, with a higher proportion of males in the PAA group than in the NSAP group (p = 0.005). Regarding clinical variables, patients with PAA showed a significantly higher prevalence of hyporexia and vomiting (p < 0.001), as well as a markedly increased prevalence of positive peritoneal irritation signs (Blumberg sign) (p < 0.001). In addition, inflammatory laboratory parameters were significantly higher in the PAA group, including total leukocyte count, ANC, ALC, SII, and C-reactive protein levels (p < 0.0001). Ultrasound-measured appendix transverse diameter was also significantly larger in patients with PAA than in those with NSAP (p < 0.0001). Comparisons of sociodemographic and clinical characteristics between patients with complete BIDIAP index data and those with missing data showed no statistically significant differences.

### Diagnostic Performance of the BIDIAP Index

Patients in the NSAP group had a median (IQR) BIDIAP index of 2 (0–3), whereas patients with acute appendicitis had substantially higher scores, with a median (IQR) of 7 (5–9) (p < 0.0001). Receiver operating characteristic analysis demonstrated excellent discriminatory performance, with an area under the curve of 0.93 (95% CI, 0.91–0.95) (Figure 1). Bootstrap resampling with 2,000 iterations yielded an AUC of 0.93 (95% CI, 0.91–0.95). Using the pre-specified cutoff of ≥ 4 points, the BIDIAP index showed a sensitivity of 90.46% and a specificity of 81.59%, with a positive likelihood ratio (LR+) of 4.91, a negative likelihood ratio (LR−) of 0.12, a positive predictive value (PPV) of 86.7%, and a negative predictive value (NPV) of 86.6%. Table 2 summarizes sensitivity, specificity, positive and negative predictive values, and likelihood ratios across the evaluated BIDIAP index cutoff points. Table 3 presents the 2×2 contingency table summarizing true positives, false positives, true negatives, and false negatives for the BIDIAP index at the prespecified cutoff of ≥ 4 points.

**Figure 1.**
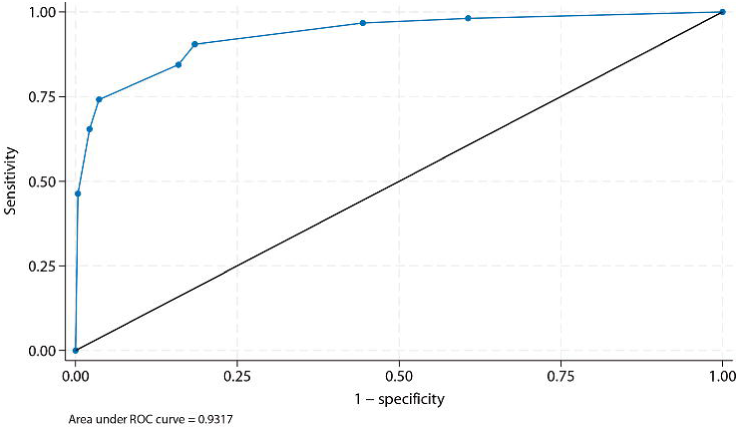
Receiver operating characteristic (ROC) curve illustrating the diagnostic performance of the BIDIAP index (PAA vs. NSAP). The BIDIAP index demonstrated excellent discrimination, with an area under the curve (AUC) of 0.93 (95% CI, 0.92–0.95).

### Calibration of the BIDIAP Index

The BIDIAP index showed excellent calibration. Logistic calibration analysis demonstrated a calibration slope of 1.00 (95% CI, 0.99–1.00) and a calibration intercept not significantly different from zero, indicating the absence of systematic over- or underestimation of risk (Figure 2). Overall predictive accuracy was high, with a Brier score of 0.10. Bootstrap internal validation using bootstrap resampling (2,000 iterations) demonstrated excellent calibration, with a calibration slope of 1.00 and an intercept not significantly different from zero, indicating stable and well-calibrated risk predictions.

**Figure 2.**
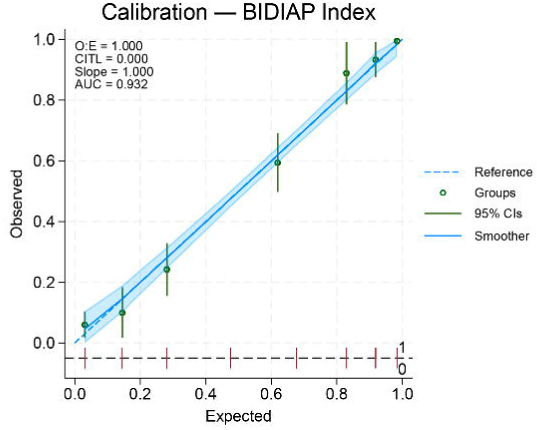
Calibration plot of the BIDIAP index showing agreement between predicted and observed probabilities of acute appendicitis. Predicted risks were grouped into deciles, with points representing observed event rates and the dashed line indicating ideal calibration. The BIDIAP index showed excellent calibration, with a calibration slope of 1.00 and an intercept close to zero.

### Decision Curve Analysis of the BIDIAP Index

Decision curve analysis demonstrated that the BIDIAP index provided a net clinical benefit across the full range of clinically relevant threshold probabilities. Throughout this interval, the net benefit of the BIDIAP index consistently exceeded those of the “treat all” and “treat none” strategies, indicating potential clinical utility across a wide range of decision thresholds (Figure 3).

**Figure 3.**
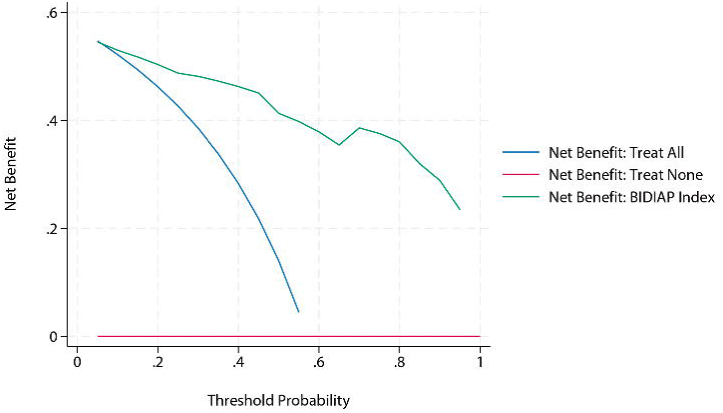
Decision curve analysis (DCA) evaluating the clinical utility of the BIDIAP index. Across the full range of threshold probabilities, the net benefit of the BIDIAP index exceeded that of both the treat-all and treat-none strategies, indicating potential clinical usefulness across a wide range of decision thresholds.

### Sensitivity Analyses

A first sensitivity analysis was performed to assess the impact of peritoneal irritation (PI) coding on the diagnostic performance of the BIDIAP index, reclassifying cases initially considered unclear as positive rather than negative for index calculation. Under this alternative assumption, the BIDIAP index maintained excellent discriminative ability, with an area under the ROC curve of 0.93 (AUC 0.93; 95% CI, 0.91–0.95). Internal validation using bootstrap resampling (2,000 iterations) confirmed the robustness of these findings, yielding a bootstrap-corrected AUC of 0.93 (95% CI, 0.91–0.95). Using the pre-specified cutoff of ≥ 4 points, the BIDIAP index showed a sensitivity of 94.01% and a specificity of 70.40%, with a positive likelihood ratio (LR+) of 3.18, and a negative likelihood ratio (LR−) of 0.09. Overall predictive accuracy remained high, with a Brier score of 0.10. Calibration remained excellent, with predicted probabilities closely aligned with the observed outcome prevalence, and DCA demonstrated unchanged clinical utility, with consistent net benefit over treat-all and treat-none strategies across the full range of clinically relevant threshold probabilities.

A second sensitivity analysis (n = 433) was performed, restricting the sample to patients in whom the appendiceal diameter was successfully measured on ultrasonography. Within this subgroup, the BIDIAP index maintained excellent discriminative performance, with an area under the receiver operating characteristic curve of 0.93. Bootstrap internal validation (2,000 resamples) confirmed the robustness of discrimination, yielding an AUC of 0.93 (95% CI, 0.91–0.96). Model calibration remained excellent in this restricted cohort, with a calibration slope of 1.00 and an intercept not significantly different from zero, indicating the absence of systematic over- or underestimation of risk. Overall predictive accuracy was high, as reflected by a low Brier score (0.09). Consistent with the primary analysis, decision curve analysis demonstrated preserved clinical utility across the full range of clinically relevant threshold probabilities, with net benefit superior to treat-all and treat-none strategies.

## Discussion

This study completes the prospective, multicenter external validation of the BIDIAP index in a real-world pediatric emergency setting. In a large, well-characterized cohort, the BIDIAP index demonstrated excellent diagnostic performance, with high discriminatory ability, as reflected by a robust area under the ROC curve, and excellent calibration, indicating accurate risk estimation across the spectrum of predicted probabilities. Moreover, DCA confirmed consistent clinical utility, with net benefit superior to treat-all and treat-none strategies across clinically relevant thresholds. Taken together, these findings support the reliability, robustness, and potential clinical value of the BIDIAP index as a simple, well-calibrated tool for assessing suspected PAA.

In relation to prior evidence, commonly used pediatric appendicitis scores, such as the PAS and pARC, have shown variable performance across external validation studies. In most published validation cohorts, reported AUC values for these tools are below 0.90 [7,9], reflecting variability across populations and clinical contexts. Moreover, many available validations have been conducted in single-center settings, which may limit generalizability. In this context, and to the best of our knowledge, the diagnostic performance observed in the present study represents one of the highest levels of discrimination reported for a clinical score undergoing prospective, multicenter external validation for the diagnosis of PAA.

An additional strength of the BIDIAP index lies in its parsimonious design. By avoiding the simultaneous inclusion of closely related hematological parameters such as leukocyte and neutrophil counts, the index reduces potential collinearity while preserving the inflammatory signal through the SII. Moreover, incorporating ultrasound appendix diameter, a radiological variable with high diagnostic yield, further enhances discriminatory performance. Importantly, non-visualization of the appendix during pediatric abdominal ultrasound, particularly in non-expert hands, is relatively common in routine clinical practice [17]. In this regard, a relevant advantage of the BIDIAP index is that reaching the prespecified diagnostic cutoff of ≥ 4 points does not require this radiological item to be positive. As demonstrated in the present validation, a combination of elevated SII (≥ 890) and clinical signs of peritoneal irritation is sufficient to identify acute appendicitis even in the absence of conclusive ultrasound findings, supporting the practical applicability of the BIDIAP index in real-world emergency settings.

From a resource-utilization and feasibility perspective, the BIDIAP index is a rapid, readily available diagnostic tool. Its calculation requires only a complete blood count (CBC), a focused physical examination, and an abdominal ultrasound, all of which are routinely available in emergency care settings. With the increasing adoption of point-of-care ultrasound (POCUS) for pediatric abdominal evaluation in emergency departments [18], combined with the low cost, rapid turnaround time, and wide availability of hematological testing, the BIDIAP index emerges as a particularly practical option. These characteristics support its potential relevance not only in high-resource environments but also in resource-limited and developing settings, where access to advanced imaging or other more complex and costly diagnostic resources, such as novel non-invasive biomarkers [19] or artificial intelligence–based models [20], may be limited, and rapid, reliable clinical decision-making is essential.

An important practical implication of our findings relates to the standardization of clinical definitions when applying the BIDIAP index. In this validation, doubtful or equivocal peritoneal irritation (PI) on physical examination was deliberately coded as absent (0 points). Although global discriminative performance, as reflected by the AUC, remained similar under alternative PI coding strategies, relevant discrepancies emerged in sensitivity and specificity at clinically relevant cutoffs. In particular, classifying equivocal PI as positive resulted in higher sensitivity but a clear loss of specificity, leading to an increased rate of false-positive classifications at the prespecified decision thresholds. From an implementation perspective, this underscores the need for explicit clinical training to emphasize that, in cases of uncertainty during abdominal examination, PI should not be scored. Overinterpretation of equivocal findings would artificially inflate the score and compromise the intended balance between sensitivity and specificity. Consistent adherence to this conservative scoring rule is therefore essential to preserve the diagnostic properties of the BIDIAP index observed in this external validation.

Lastly, regarding cutoffs, a value of ≥ 4 points was selected a priori as the primary threshold because it was identified as the optimal cutoff in the original derivation cohort and therefore was prespecified for external validation. Nevertheless, alternative cutoffs of the BIDIAP index offer clinically meaningful information depending on the intended use of the index. At the ≥ 4 threshold, the index achieves high overall discrimination and specificity, making it well-suited for identifying children at high risk of PAA. However, with a sensitivity of approximately 90% and a negative likelihood ratio (LR−) of 0.12, this cutoff may be suboptimal for safe rule-out in emergency settings, where LR− values below 0.1 are generally preferred to confidently exclude serious pathology without further testing. In this context, lower thresholds merit consideration. Notably, a cutoff of ≥ 2 points yields a very high sensitivity (98%) and an excellent LR− of 0.07, supporting its potential use as a screening or rule-out threshold, albeit at the cost of reduced specificity. These findings suggest that a dual-threshold strategy, with a low cutoff optimized to rule out PAA and a higher cutoff optimized to rule in PAA, may enhance clinical decision-making. Such an approach is analogous to strategies commonly applied with established scores, such as the Alvarado score. It may better reflect the sequential, risk-stratified nature of decision-making in pediatric emergency care.

This study has several important strengths. It was conducted using a prospective, methodologically rigorous design, with consecutive patient enrollment across multiple tertiary centers, thereby enhancing internal validity and clinical relevance. The statistical methodology was comprehensive and appropriately aligned with the data’s nature, incorporating discrimination, calibration, internal validation, and decision curve analysis. In addition, the study adhered to established reporting standards for diagnostic and prognostic research, including the STARD and TRIPOD recommendations, further supporting transparency and methodological robustness. Lastly, the final sample markedly exceeded the minimum sample size required for external validation, resulting in narrower uncertainty around discrimination and calibration estimates.

Some limitations should also be acknowledged. Although multicenter, the study was conducted within a single country, and the performance of the BIDIAP index may differ in other geographic, sociodemographic, or healthcare settings. Nevertheless, the participating centers spanned different regions of Spain (Madrid, Castilla y León, and Aragón), providing some geographic and population variability. In addition, data required to compute the BIDIAP index according to the prespecified operational criteria were available for 644 of 719 eligible patients. Although no significant differences in baseline characteristics were observed, the possibility of unmeasured confounding due to missing ultrasonographic or clinical documentation cannot be entirely ruled out.

A further limitation relates to the management of cases in which the appendix was not visualized on ultrasonography. In these patients, the absence of a measurable appendiceal caliber was operationally interpreted as the absence of pathological enlargement, corresponding to 0 points for the appendiceal diameter component of the BIDIAP index, which is the highest-weighted item (4 points). It should be acknowledged that, in routine clinical practice, non-visualization of the appendix does not necessarily imply a completely normal examination. Indirect ultrasonographic signs, such as hyperechogenic periappendiceal fat or the presence of free fluid in the right iliac fossa, may still raise suspicion of PAA despite the appendix itself not being identified. However, by the definition of the scoring system, all such cases were classified as negative for the appendiceal diameter item. Consequently, these patients could only reach the prespecified diagnostic cutoff of ≥ 4 points if the remaining two components of the index, namely an elevated SII (≥ 890) and the presence of peritoneal irritation, were simultaneously present. This approach reflects common clinical practice in pediatric emergency ultrasound, where non-visualization of the appendix, particularly in low-risk settings, is frequently considered a reassuring finding rather than an indeterminate one. Nevertheless, from a statistical perspective, this assumption may not always hold, as non-visualization can also be influenced by factors such as patient body mass index [21], bowel gas, or operator-dependent limitations of the acoustic window. Accordingly, while this strategy is clinically defensible and consistent with the intended real-world applicability of the BIDIAP index, it may lead to misclassification in a subset of patients and could influence diagnostic performance estimates. Notably, however, sensitivity analyses specifically addressing this issue confirmed that the diagnostic performance of the BIDIAP index remained excellent and, in analyses restricted to patients with a measured appendiceal diameter, was even slightly improved, thereby supporting the robustness of the model under alternative, clinically plausible assumptions.

In conclusion, in this prospective, multicenter external validation study, the BIDIAP index demonstrated excellent diagnostic performance for pediatric acute appendicitis, combining strong discrimination, accurate calibration, and consistent clinical utility on decision curve analysis. Its parsimonious design, reliance on widely available clinical, laboratory, and ultrasound variables, and robustness across sensitivity analyses support its practicality in routine emergency settings. Together, these findings suggest that the BIDIAP index is a reliable and clinically applicable tool for evaluating suspected pediatric acute appendicitis, with potential usefulness across a wide range of healthcare settings.

## CRediT authorship contribution statement

**JAM:** Conceptualization and study design; literature search and selection; data curation and extraction; formal analysis; investigation; methodology; project administration; resources; validation; visualization; writing – original draft; writing – review and editing.

**RFA, CDM**: Data curation and extraction; resources; validation; visualization; writing – review and editing.

**AHM, AEV, MIO, LFR, PAR, RD, LRV, MLO, MCC, CRM, SSA:** Data curation and extraction; resources; writing – review and editing.

## ETHICAL APPROVAL

The Institutional Review Board of Complejo Asistencial Universitario de León reviewed and approved this study under code 24035. The Institutional Review Board of Hospital Fundación Jiménez Díaz reviewed and approved this study under code 20324. All other participating centers formally acknowledged, validated, and adhered to the Institutional Review Board approval granted by the Complejo Asistencial Universitario de León. Before submitting this article, verbal and written informed consent were obtained from the patients’ legal guardians. This study was conducted in accordance with the principles of the Helsinki Declaration (2013 statement).

## STATEMENT OF AVAILABILITY OF THE DATA USED DURING THE STUDY

The data used to carry out this study are available upon reasonable request from the corresponding author.

## References

[1]. Waseem M, Wang CF. Pediatric appendicitis. In: StatPearls [Internet]. Treasure Island (FL): StatPearls Publishing; 2025 Jan–. 2025 Jun 17 [Accessed 2025 Dec 29]. Available from: StatPearls Publishing. PMID: 28722894.

[2]. Michelson KA, Reeves SD, Grubenhoff JA, Cruz AT, Chaudhari PP, Dart AH, Finkelstein JA, Bachur RG. Clinical Features and Preventability of Delayed Diagnosis of Pediatric Appendicitis. JAMA Netw Open. 2021 Aug 2;4(8):e2122248. doi: 10.1001/jamanetworkopen.2021.22248. PMID: 34463745; PMCID: PMC8408667.

[3]. Alvarado A. A practical score for the early diagnosis of acute appendicitis. Ann Emerg Med. 1986 May;15(5):557–64. doi: 10.1016/s0196-0644(86)80993-3. PMID: 3963537.

[4]. Pogorelić Z, Rak S, Mrklić I, Jurić I. Prospective validation of Alvarado score and Pediatric Appendicitis Score for the diagnosis of acute appendicitis in children. Pediatr Emerg Care. 2015 Mar;31(3):164–8. doi: 10.1097/PEC.0000000000000375. PMID: 25706925.

[5]. Bai S, Hu S, Zhang Y, Guo S, Zhu R, Zeng J. The Value of the Alvarado Score for the Diagnosis of Acute Appendicitis in Children: A Systematic Review and Meta-Analysis. J Pediatr Surg. 2023 Oct;58(10):1886–1892. doi: 10.1016/j.jpedsurg.2023.02.060. Epub 2023 Mar 6. PMID: 36966018.

[6]. Samuel M. Pediatric appendicitis score. J Pediatr Surg. 2002 Jun;37(6):877–81. doi: 10.1053/jpsu.2002.32893. PMID: 12037754.

[7]. Bhatt M, Joseph L, Ducharme FM, Dougherty G, McGillivray D. Prospective validation of the pediatric appendicitis score in a Canadian pediatric emergency department. Acad Emerg Med. 2009 Jul;16(7):591–6. doi: 10.1111/j.1553-2712.2009.00445.x. Epub 2009 Jun 22. PMID: 19549016.

[8]. Kharbanda AB, Vazquez-Benitez G, Ballard DW, Vinson DR, Chettipally UK, Kene MV, Dehmer SP, Bachur RG, Dayan PS, Kuppermann N, O’Connor PJ, Kharbanda EO. Development and Validation of a Novel Pediatric Appendicitis Risk Calculator (pARC). Pediatrics. 2018 Apr;141(4):e20172699. doi: 10.1542/peds.2017-2699. Epub 2018 Mar 13. PMID: 29535251; PMCID: PMC5869337.

[9]. Cotton DM, Vinson DR, Vazquez-Benitez G, Margaret Warton E, Reed ME, Chettipally UK, Kene MV, Lin JS, Mark DG, Sax DR, McLachlan ID, Rauchwerger AS, Simon LE, Kharbanda AB, Kharbanda EO, Ballard DW; Clinical Research on Emergency Services and Treatments (CREST) Network. Validation of the Pediatric Appendicitis Risk Calculator (pARC) in a Community Emergency Department Setting. Ann Emerg Med. 2019 Oct;74(4):471–480. doi: 10.1016/j.annemergmed.2019.04.023. Epub 2019 Jun 19. PMID: 31229394; PMCID: PMC8364751.

[10]. Michelson KA, Bachur RG, Rangel SJ, Finkelstein JA, Monuteaux MC, Goyal MK. Disparities in Diagnostic Timeliness and Outcomes of Pediatric Appendicitis. JAMA Netw Open. 2024 Jan 2;7(1):e2353667. doi: 10.1001/jamanetworkopen.2023.53667. PMID: 38270955; PMCID: PMC10811560.

[11]. Arredondo Montero J, Bardají Pascual C, Antona G, Ros Briones R, López-Andrés N, Martín-Calvo N. The BIDIAP index: a clinical, analytical and ultrasonographic score for the diagnosis of acute appendicitis in children. Pediatr Surg Int. 2023 Apr 10;39(1):175. doi: 10.1007/s00383-023-05463-5. PMID: 37038002; PMCID: PMC10085908.

[12]. Cohen JF, Korevaar DA, Altman DG, Bruns DE, Gatsonis CA, Hooft L, Irwig L, Levine D, Reitsma JB, de Vet HC, Bossuyt PM. STARD 2015 guidelines for reporting diagnostic accuracy studies: explanation and elaboration. BMJ Open. 2016 Nov 14;6(11):e012799. doi: 10.1136/bmjopen-2016-012799. PMID: 28137831; PMCID: PMC5128957.

[13]. Collins GS, Moons KGM, Dhiman P, Riley RD, Beam AL, Van Calster B, Ghassemi M, Liu X, Reitsma JB, van Smeden M, Boulesteix AL, Camaradou JC, Celi LA, Denaxas S, Denniston AK, Glocker B, Golub RM, Harvey H, Heinze G, Hoffman MM, Kengne AP, Lam E, Lee N, Loder EW, Maier-Hein L, Mateen BA, McCradden MD, Oakden-Rayner L, Ordish J, Parnell R, Rose S, Singh K, Wynants L, Logullo P. TRIPOD+AI statement: updated guidance for reporting clinical prediction models that use regression or machine learning methods. BMJ. 2024 Apr 16;385:e078378. doi: 10.1136/bmj-2023-078378. Erratum in: BMJ. 2024 Apr 18;385:q902. doi: 10.1136/bmj.q902. PMID: 38626948; PMCID: PMC11019967.

[14]. Hanley JA, McNeil BJ. The meaning and use of the area under a receiver operating characteristic (ROC) curve. Radiology. 1982 Apr;143(1):29–36. doi: 10.1148/radiology.143.1.7063747. PMID: 7063747.

[15]. Hanley JA, McNeil BJ. A method of comparing the areas under receiver operating characteristic curves derived from the same cases. Radiology. 1983 Sep;148(3):839–43. doi: 10.1148/radiology.148.3.6878708. PMID: 6878708.

[16]. Vickers AJ, Elkin EB. Decision curve analysis: a novel method for evaluating prediction models. Med Decis Making. 2006 Nov-Dec;26(6):565–74. doi: 10.1177/0272989X06295361. PMID: 17099194; PMCID: PMC2577036.

[17]. Mittal MK, Dayan PS, Macias CG, Bachur RG, Bennett J, Dudley NC, Bajaj L, Sinclair K, Stevenson MD, Kharbanda AB; Pediatric Emergency Medicine Collaborative Research Committee of the American Academy of Pediatrics. Performance of ultrasound in the diagnosis of appendicitis in children in a multicenter cohort. Acad Emerg Med. 2013 Jul;20(7):697–702. doi: 10.1111/acem.12161. PMID: 23859583; PMCID: PMC5562364.

[18]. Balbo S, Pini CM, Raffaldi I, Delmonaco AG, Castagno E, Guanà R, Di Rosa G, Bondone C. Accuracy of point-of-care ultrasound in the diagnosis of acute appendicitis in a pediatric emergency department. J Clin Ultrasound. 2024 Jun;52(5):485–490. doi: 10.1002/jcu.23658. Epub 2024 Mar 4. PMID: 38436504.

[19]. Arredondo Montero J, Ros Briones R, Fernández-Celis A, López-Andrés N, Martín-Calvo N. Diagnostic Performance of Serum Leucine-Rich Alpha-2-Glycoprotein 1 in Pediatric Acute Appendicitis: A Prospective Validation Study. Biomedicines. 2024 Aug 11;12(8):1821. doi: 10.3390/biomedicines12081821. PMID: 39200285; PMCID: PMC11352011.

[20]. Chekmeyan M, Liu SH. Artificial intelligence for the diagnosis of pediatric appendicitis: A systematic review. Am J Emerg Med. 2025 Jun;92:18–31. doi: 10.1016/j.ajem.2025.02.023. Epub 2025 Feb 17. PMID: 40048888.

[21]. Pfeifer CM, Xie L, Atem FD, Mathew MS, Schiess DM, Messiah SE. Body mass index as a predictor of sonographic visualization of the pediatric appendix. Pediatr Radiol. 2022 Jan;52(1):42–49. doi: 10.1007/s00247-021-05176-8. Epub 2021 Sep 15. PMID: 34524472.

