## Supplementary File 3 for "Prospective Multicenter External Validation of the BIDIAP Index for the Diagnosis of Pediatric Acute Appendicitis"

**Inclusion Criteria**

The study will include patients aged 0 to 18 years (inclusive) presenting to the Pediatric Emergency department of any participating center with acute abdominal pain suggestive of acute appendicitis, defined as pain initially located in the mesogastrium with subsequent migration to the right lower quadrant, or pain with onset in the right lower quadrant. Patients must present with at least one of the following associated symptoms: anorexia, nausea, vomiting, low-grade fever, fever, or diarrhea.

The histopathological diagnosis of the surgical specimen will confirm inclusion in the case group. Patients in whom the histopathological examination shows no findings compatible with acute appendicitis (“negative appendectomy”) will be classified within the control group (operated patients without a diagnosis of appendicitis).

A control group will be established, with a target case-to-control ratio of 1:1.The control group will consist of patients aged 0 to 18 years (inclusive) presenting to the pediatric emergency department of any participating center with acute abdominal pain suggestive of acute appendicitis, defined as pain initially located in the mesogastrium with subsequent migration to the right lower quadrant, or pain with onset in the right lower quadrant. Patients must present with at least one of the following associated symptoms: anorexia, nausea, vomiting, low-grade fever, fever, or diarrhea. In these patients, acute appendicitis must be ruled out based on the identification of an alternative, more probable diagnosis and a favorable spontaneous clinical course, leading to discharge home. In this control group, a structured telephone follow-up will be conducted 2–3 weeks after the emergency department visit to confirm favorable clinical evolution and the absence of signs or symptoms suggestive of acute appendicitis after discharge.

After written informed consent has been obtained, patients will be included in the study.

**Exclusion Criteria**

- Patients with a clear clinical diagnosis of acute appendicitis and clinical instability requiring immediate surgical intervention without prior complementary diagnostic testing.
- Patients with metastatic neoplastic disease.
- Patients with hematological disorders.
- Patients with active autoimmune diseases.
- Patients with a previous appendectomy.
- Patients treated with immunosuppressive therapy within the 28 days before Emergency Department evaluation.
- Patients treated with systemic corticosteroids within the 14 days before Emergency Department evaluation.
- Patients with a history of abdominal trauma before Emergency Department evaluation.
- Patients with invasive abdominal neoplasms.
- Patients who underwent abdominal radiological imaging within the 14 days before Emergency Department evaluation (except for plain abdominal radiography).
- Explicit refusal of study participation by the patient’s parents or legal guardians.
