## Supplementary File 4 for "Prospective Multicenter External Validation of the BIDIAP Index for the Diagnosis of Pediatric Acute Appendicitis"

Potentially eligible participants n=725

Index test positive (n=383)

Referece standard (n=383)

Final diagnosis Target condition present (n=332) Target condition absent (n=51) Inconclusive (n=0)

Index test negative (n=261)

Reference standard (n=261)

Final diagnosis Target condition present (n=35) Target condition absent (n=226) Inconclusive (n=0)

Index test inconclusive (n=0)

Excluded participants n=6 -Reason 1 (n=1): duplicate entry -Reason 2 (n=3): no informed consent -Reason 3 (n=1): wrong inclusion criteria Reason 4 (n=1): no standard reference

Index test n=644
