## Supplementary material for "Prospective Multicenter External Validation of the BIDIAP Index for the Diagnosis of Pediatric Acute Appendicitis": Table 1

Table 1. Sociodemographic and clinical characteristics of the patients.

| Sociodemographics | Group 1 (NSAP)  *n* = 305 | Group 2 (PAA)  *n* = 414 | *P*-value |
| --- | --- | --- | --- |
| Age, y | 10.0 (2.87) | 10.13 (3.1) | 0.59* |
| Sex, M/F | 156/158 | 243/161 | 0.005** |
| Duration of abdominal pain, h^1^ | 24 (12-48) | 24 (12-48) | 0.22*** |
| Hyporexia, Yes/No | 149/157 | 250/146 | <0.001** |
| Fever at home (>37.8 °C), Yes/No | 88/224 | 118/284 | 0.74** |
| Vomiting, Yes/No | 121/192 | 229/172 | <0.001** |
| Diarrhea, Yes/No | 60/253 | 75/327 | 0.81** |
| Peritoneal irritation (Blumberg sign), Yes/No/Unclear | 85/108/86 | 249/57/67 | <0.001** |
| Absolute leukocyte count (1x10^9^/L)^1^ | 9.1 (6.9-13.2) | 15.5 (12.4-18.4) | <0.0001*** |
| Absolute lymphocyte count (1x10^9^/L)^1^ | 2.13 (1.38-2.9) | 1.6 (1.08-2.13) | <0.0001*** |
| Absolute neutrophil count (1x10^9^/L)^1^ | 5.55 (3.89-10) | 12.6 (9-15.6) | <0.0001*** |
| Absolute platelet count (1x10^9^/L) | 290.15 (74.72) | 297.57 (73.56) | 0.19* |
| SII^1^ | 759.33 (410.03-1721) | 2383.24 (1239.5-4114.53) | <0.0001*** |
| Appendiceal ultrasonographic transverse diameter, mm^1^ | 5 (4-6.25) | 9 (7.5-10) | <0.0001*** |
| C-Reactive Protein (mg/L)^1^ | 5 (0.6-21.5) | 24.8 (8-64.7) | <0.0001*** |
| BIDIAP score^1^ | 2 (0-3) | 7 (5-9) | <0.0001*** |

*:t-test (two-sided); **:χ² test (two-sided); ***: Mann–Whitney U test (two-sided)

1: Median (interquartile range)

Numbers are the mean (standard deviation) or the total number.

*NSAP* non-surgical abdominal pain, *PAA* pediatric acute appendicitis, *M* male, *y* years, *h* hours, *SII* Systemic-Immune Inflammation Index,

*Denominators vary across variables due to missing data.*
