## Supplementary material for "Prospective Multicenter External Validation of the BIDIAP Index for the Diagnosis of Pediatric Acute Appendicitis": Table 2

Table 2. Cutoffs for BIDIAP Index

Pediatric acute appendicitis (PAA) vs non-surgical abdominal pain (NSAP)

| BIDIAP Index cut-off value | Correctly classified (%) | Sensitivity (%) | Specificity (%) | LR+ | LR- | PPV (%) | NPV (%) |
| --- | --- | --- | --- | --- | --- | --- | --- |
| ≥2 | 72.8 | 98.1 | 39.4 | 1.62 | 0.05 | 68.2 | 94.0 |
| ≥3 | 79.0 | 96.7 | 55.6 | 2.18 | 0.06 | 74.3 | 92.8 |
| **≥4** | **86.7** | **90.5** | **81.6** | **4.91** | **0.12** | **86.7** | **86.6** |
| ≥5 | 84.3 | 84.5 | 84.1 | 5.32 | 0.19 | 87.6 | 80.3 |
| ≥6 | 83.7 | 74.1 | 96.4 | 20.5 | 0.27 | 96.5 | 73.8 |
| ≥7 | 79.4 | 65.4 | 97.8 | 30.2 | 0.35 | 97.6 | 68.1 |

*LR* Likelihood ratio; *NPV* Negative predictive value, *PPV* Positive predictive value
