## Supplementary material for "Prospective Multicenter External Validation of the BIDIAP Index for the Diagnosis of Pediatric Acute Appendicitis": Table 3

Table 3. Contingency Table (True Positives, False Positives, True Negatives, and False Negatives) for the BIDIAP Index at the Prespecified Cutoff of ≥ 4 Points

Pediatric acute appendicitis (PAA) vs non-surgical abdominal pain (NSAP)

| BIDIAP Index cut-off value | PAA | NSAP | Total |
| --- | --- | --- | --- |
| ≥4 | 332 (TP) | 51 (FP) | 383 |
| <4 | 35 (FN) | 226 (TN) | 261 |
| Total | 367 | 277 | 644 |

*TP* True positives; *TN* True negatives, *FP* False positives; *FN* False negatives
